# Estimating missing deaths in Delhi’s COVID-19 data

**DOI:** 10.1101/2020.07.29.20164392

**Authors:** Shoibal Chakravarty

## Abstract

A sero-prevalence survey in Delhi measured an infection rate of 23.48% and an implied infection fatality rate (IFR) of 0.06%. Modeling using age group based IFRs from France, Spain and Lombardia project an average IFR that is significantly higher than currently estimated. We show that at least 1500-2500 COVID-19 deaths in the 60+ age group are missing.

---

A recent sero-prevalence survey (PTI 2020) found that 23.48% of Delhi has antibodies for SARS-CoV-2, the virus that causes COVID-19. This survey was conducted between June 27 and July 10. As the time to develop antibodies from date of infection is roughly the same as the the time to death for severe cases, we can estimate that Delhi’s infection fatality rate (IFR) to be 0.06% using Delhi’s COVID-19 death tally on the 4th of July (3004)(COVID19INDIA 2020) and an estimated population of 20.2 million (MoHFW 2019) in 2020. This rate is remarkably low compared to the estimate of 0.68% (0.53%-0.82%) from a systematic review (Meyerowitz-Katz and Merone 2020) of research on COVID-19 IFRs. Good quality data that might help us understand the reason behind this low IFR has been difficult to find. I try to resolve this puzzle by piecing together publicly available data, and conclude that the COVID-19 records are missing a majority of the deaths in the 60+ age group.

## Estimating Delhi’s IFR

COVID-19 causes complications leading to hospitalization and death in a small fraction of the infected. These are mostly the elderly with comorbidities like hyper-tension, diabetes and respiratory disorders. The IFR in a country or region depends strongly on the share of the elderly, the comorbidities and disease-profile, and the capacity of the health system to respond to a surge in demand for critical care and hospitalization.

India is a relatively young country and the IFR is expected to be lower than countries with older populations like Spain, France or Italy. A country wide infection fatality rate comparison between countries with very different population structures like India or France wouldn’t make sense but comparing the fatality rates in similar age groups would be appropriate. For example, a comprehensive study (Salje et al. 2020) of France’s COVID-19 outbreak estimated these age group based IFRs (with error ranges).

The IFR goes up rapidly with age, especially past the age of 50. India’s significantly younger population would result in a lower average IFR compared to France. In Figure 1 I use a recent Census population projection (MoHFW 2019) to estimate the 2020 Delhi population distribution in different age groups. I will use both 2011 and the estimated 2020 distributions as the Delhi of 2020 is older (share of people in the 80+ group nearly doubles), and this makes a big difference to COVID-19 fatalities.

**Figure 1:**
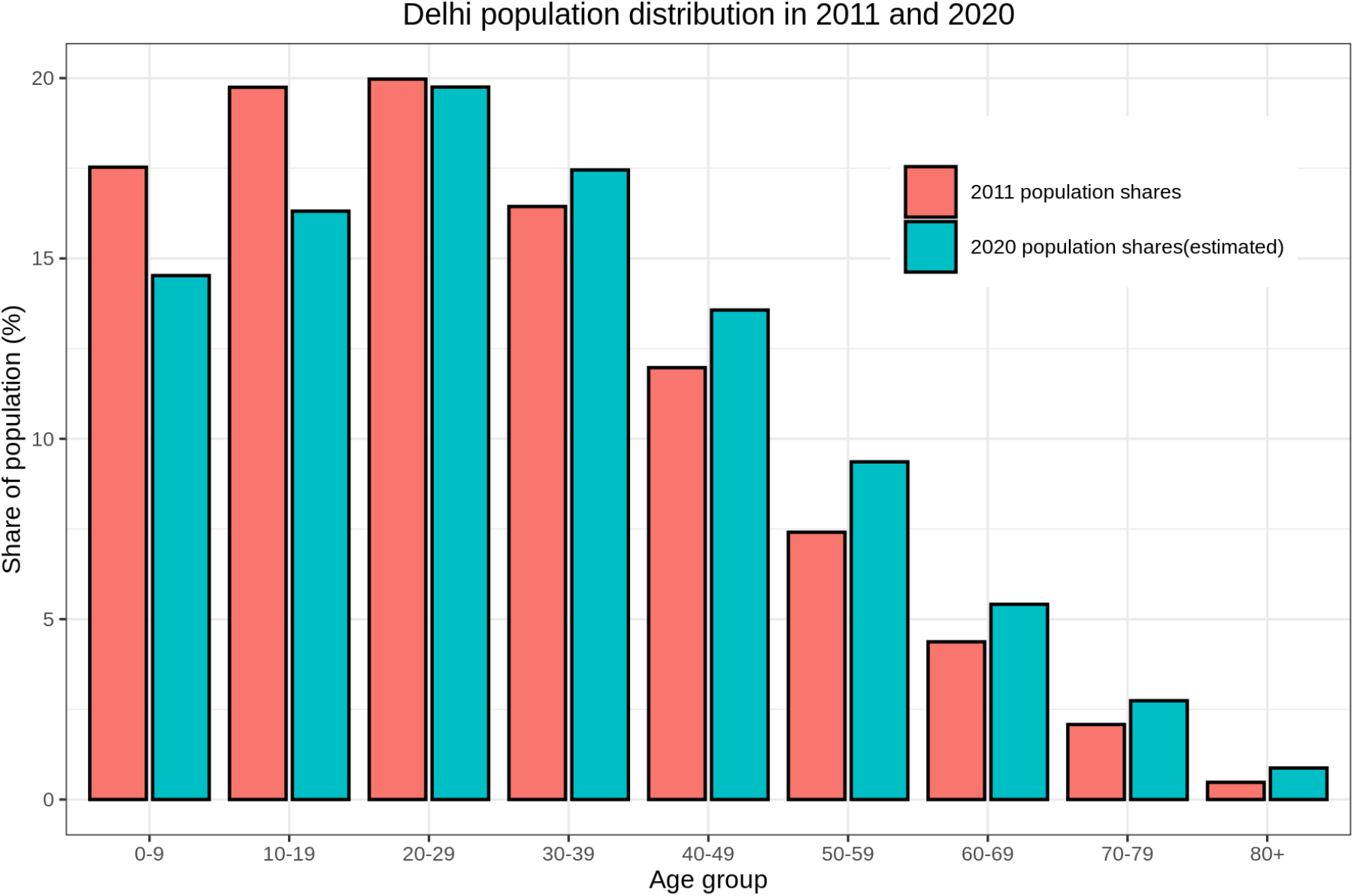
Delhi’s population distribution by 10 year age groups in 2011 compared to the projected distribution in 2020. The increase in share of people in the 60+ age bracket will play an important role in Delhi’s infection fatality rate as COVID-19.

I use age based infection rates of China (Verity et al. 2020), France (Salje et al. 2020), Lombardia (Italy) (Modi et al. 2020), New York (Yang et al. 2020) and Spain (Bevand [2020] 2020) as models to estimate Delhi COVID-19 deaths for a sero-positivity of 23.48%. I assume that all age groups are uniformly infected as age based infection rates were not released. As Figure 2 shows, Delhi’s IFR turn out to be significantly lower than what we would expect if the age group based IFRs of any of the five models regions considered above. For example, Delhi’s average IFR using France’s numbers are less than half of France’s in Table 1. Delhi’s COVID-19 deaths are anywhere between one-half to one-tenth of what the various model regions suggest, and lower than any lower error bound as well! We also see that using the estimated 2020 population distribution instead of 2011 would lead to significantly higher number of projected COVID-19 deaths.

**Table 1:** Age groups wise IFRs in France

| Age group | Infection fatality rate in % |
| --- | --- |
| <20 | 0.001 (0-0.002) |
| 20-29 | 0.005 (0.003-0.01) |
| 30-39 | 0.02 (0.01-0.03) |
| 40-49 | 0.05 (0.03-0.08) |
| 50-59 | 0.2 (0.1-0.3) |
| 60-69 | 0.7 (0.4-1.2) |
| 70-79 | 1.9 (1.1-3.2) |
| 80+ | 8.3 (4.7-13.5) |
| Average | 0.5 (0.3-0.9) |

**Figure 2:**
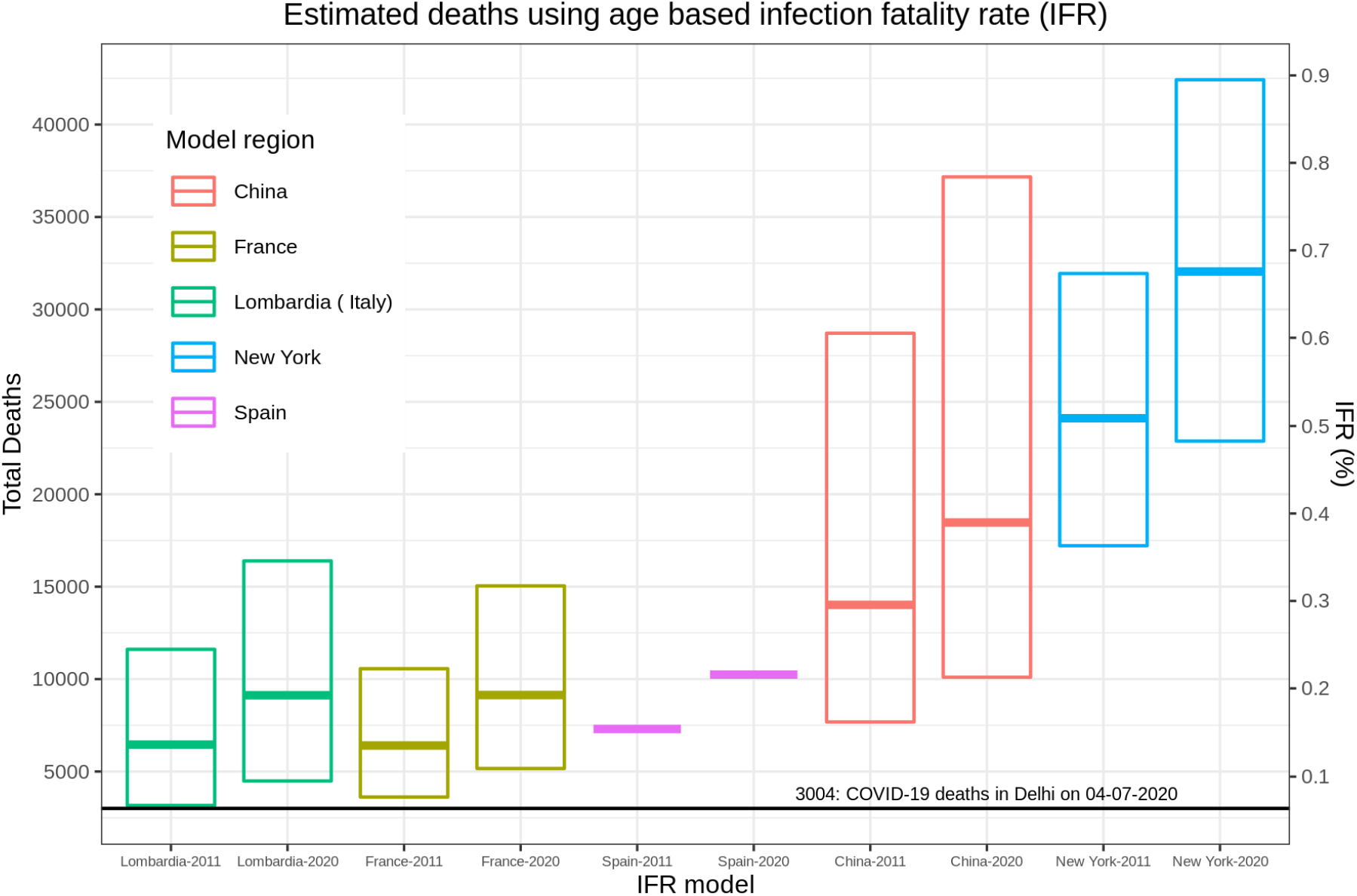
Delhi had 3004 officially confirmed COVID-19 deaths on 4th July, 2020, leading to an infection fatality rate (IFR) of 0.06% based on a seroprevalence of 23.48%. This is exceptionally low compared to the expected number of deaths (and IFR) if we apply the age-based infection fatality rates of countries and regions that have had a large COVID-19 outbreak.

## Estimating missing deaths

Delhi’s lower than expected IFR is surprising as there is no good reason to expect that an Indian in a particular age group is healthier, have better immunity, or access to better health care than a person in Europe in the same age group. An age group wise infection fatality rate of Delhi and the other models regions can shed some light on this puzzle. Unfortunately, it seems that the last time Delhi release any data of deaths by age group was on the 20th of May (DHGS Delhi 2020) when Delhi had 176 COVID-19 deaths only. Many cities and states have been more forthcoming with such data. In Figure 3, I plot the cumulative share of deaths vs. age groups for nation-wide COVID-19 data from the Integrated Disease Surveillance Programme (Ghosh 2020), Mumbai (BMC 2020), Bengaluru (BBMP 2020) and Surat (SMC 2020) on the same graph. The distribution of deaths by age groups in large urban areas is remarkably similar, perhaps reflecting their similar age and comorbidity profiles. Cities like Kolkata and states like Kerala are likely to have older populations but the differences are likely to be small. Thus far, about 60% of COVID-19 deaths have been recorded in the 10 largest metros so the IDSP data also has a similar shape. Given the absence of a better alternative, I will use the Mumbai age group wise death distribution data to model Delhi’s death distribution.

**Figure 3:**
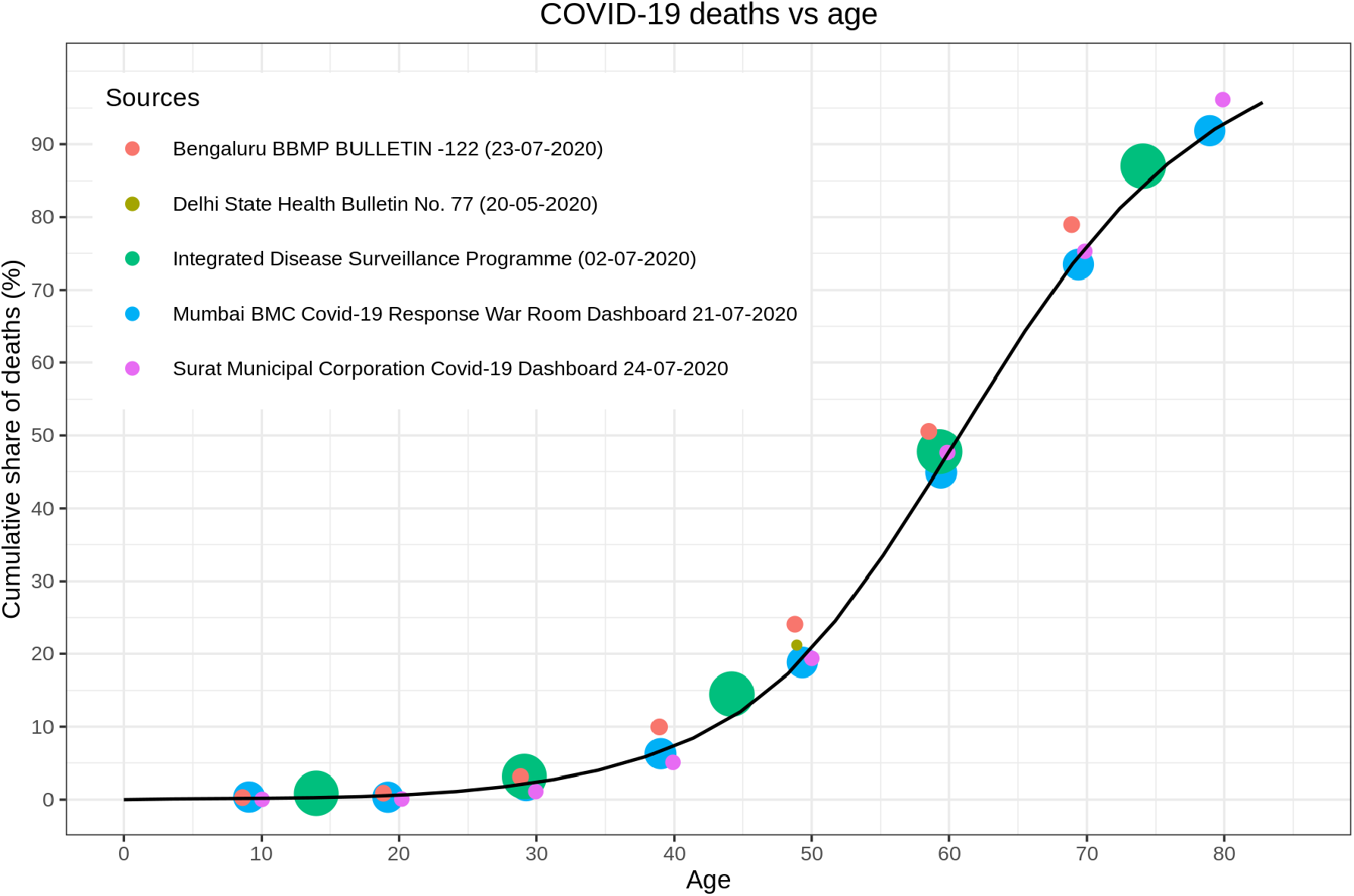
The distribution of deaths by age groups seems to be very similar in large urban areas like Mumbai, Bengaluru and Surat, and compare well with the data release by the Integrated Disease Surveillance Programme. (The size of the circles are proportional to the total number of deaths in the respective reports.)

I distribute Delhi’s COVID-19 deaths in different age groups using Mumbai as a model, and compare with the expected share of deaths in different age groups that one would expect if the age wise IFRs of France, Italy etc are applied. Figure 4 shows the histogram of these expected death shares. Firstly, note that black error bars on the histogram that shows the higher deaths in older age groups if the 2011 population distribution is replaced with the 2020 distribution. The most striking difference is that Delhi’s has a significantly higher share of deaths in relatively younger age groups (50-69 years). If higher prevalence of comorbidities are the reason for higher than expected deaths in the 50-69 age groups, the same argument should apply (perhaps, more so) for older age groups. The relative drop in share of deaths in the 70+ age groups is unexpected.

**Figure 4:**
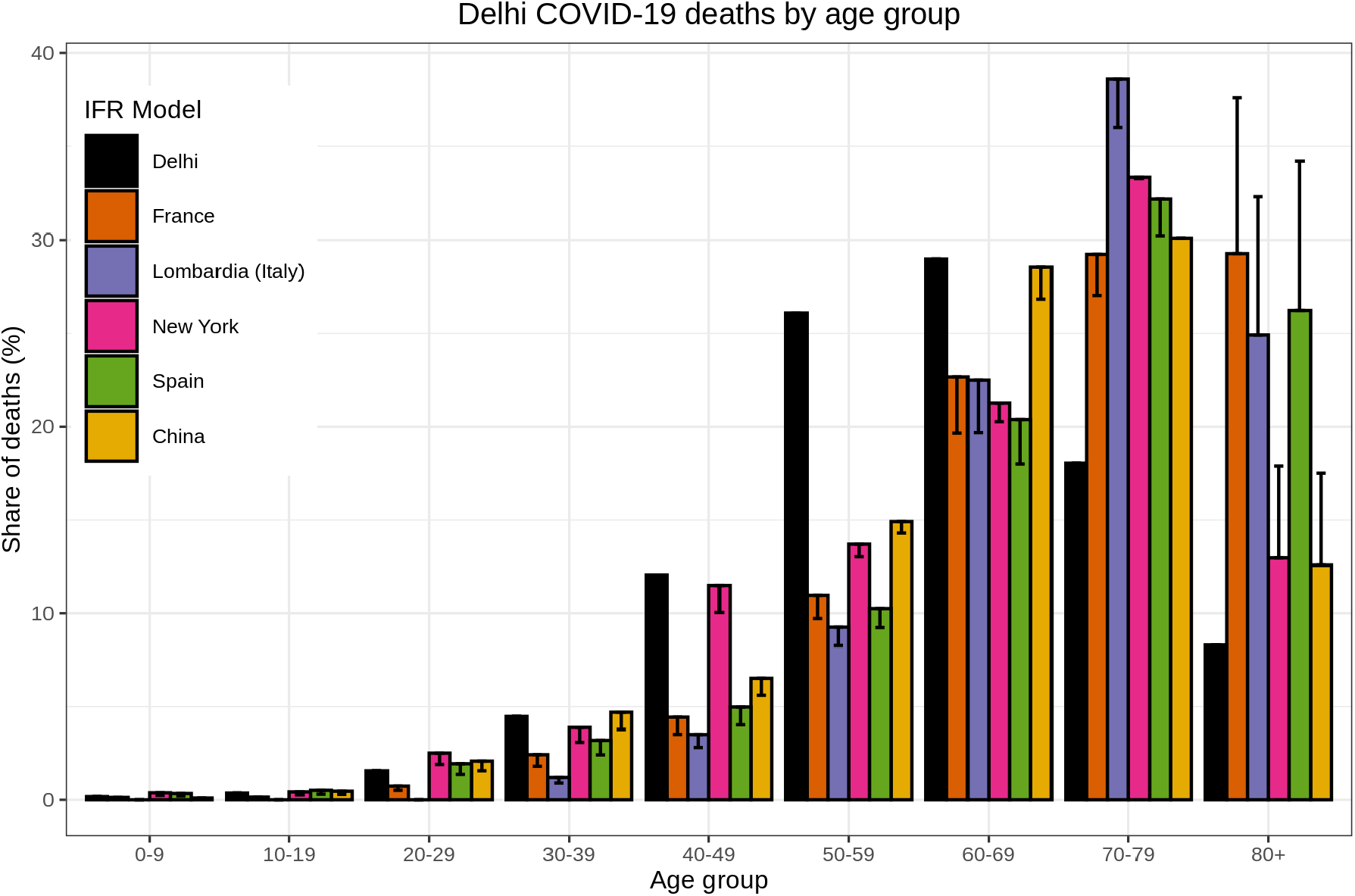
The estimated share of Delhi’s COVID-19 deaths in different age groups differs significantly from the distribution we would expect if we applied the age-based infection fatality rates of countries and regions that have had a large COVID-19 outbreak. The error bars show the increased share of deaths in the 80+ age group if we use the 2020 population distribution instead of the 2011 distribution. Only 26% of the deaths are in the 70+ age group though 58% (39%-69%) are expected. On the other hand, 55% of Delhi’s deaths are in the 50-69 age group though only 33% (27%-46%) are expected.

Let us compare the estimated number of deaths in different age groups (distributed according to Mumbai’s death distribution) with the expected number of deaths in different age groups if the age based IFRs of the European countries are applied. The China and New York IFRs are higher for all age groups and give significantly higher deaths and I’ll drop these models from the rest of the analysis. If I compare Delhi’s estimated death distribution with the expected deaths from the Europe IFR models (brown and purple dots and lines in Figure 5) I find that there is fairly good agreement for lower (30-59) age groups but very significant under-counting of deaths in the higher (60+) age groups. Conversely, if I compare Delhi’s deaths with the “low IFR” models (created using the lower error bounds of the IFR estimates), Delhi has significantly higher than expected deaths in the lower (30-59) age groups but lower deaths in the 70+ age groups. If I assume that Delhi’s COVID-19 death statistics are good for those below 60 years of age, this analysis projects that at least 1500-2500 deaths in the 60+ age groups are missing from the records.

**Figure 5:**
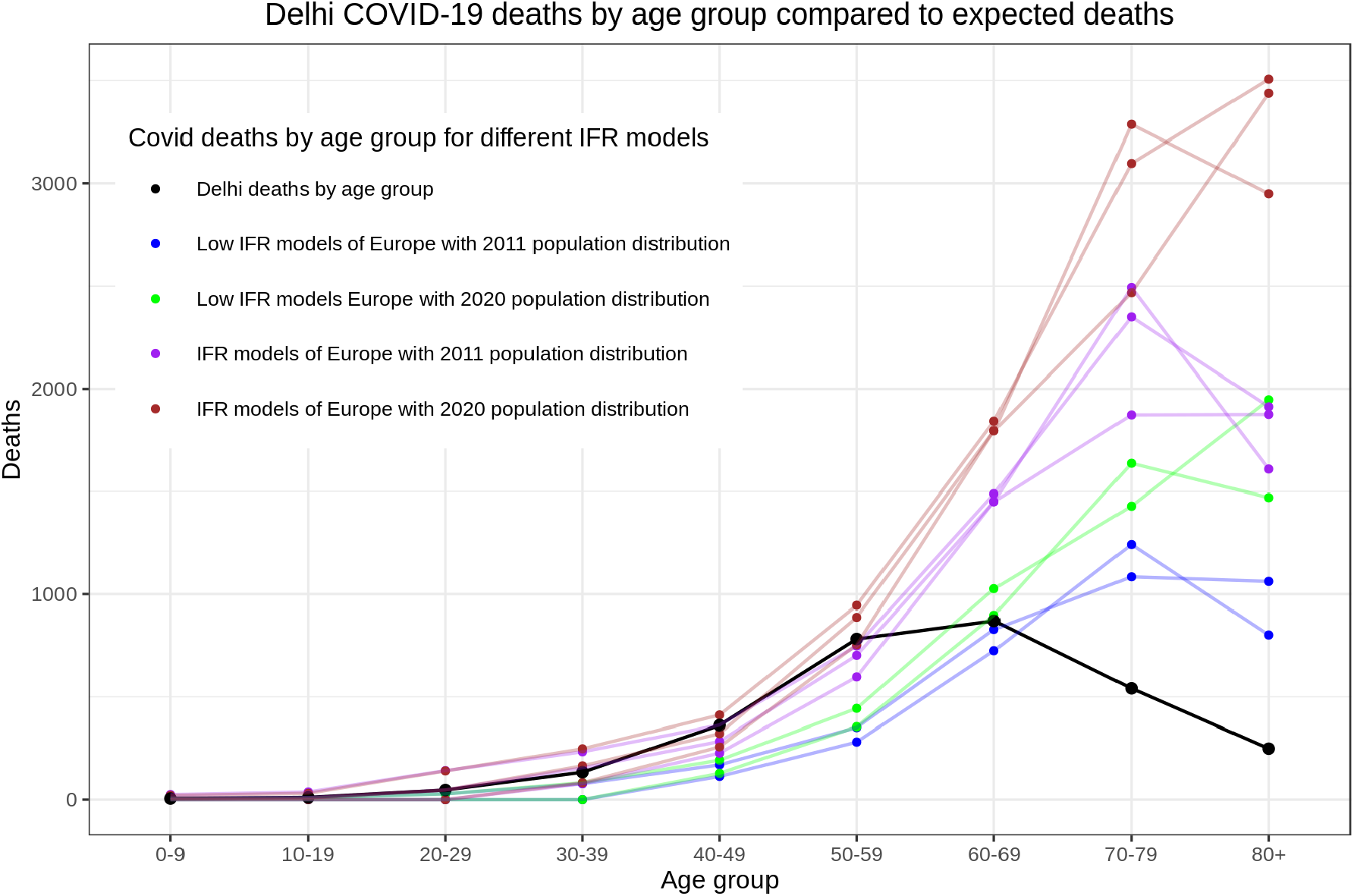
Delhi’s death distribution by age groups show a surprising divergence from the estimates we expect if we apply the IFRs of European countries/regions like Spain, France and Lombardia. The “low” numbers above are for the lower error bounds of the age based IFR estimates. Note the significant increase in expected deaths in the 70+ age groups when we use the 2020 population distribution instead of 2011. Even if we assume that Delhi’s COVID-19 death records are fairly accurate for those below 60 years, this analysis suggests that at least 500 deaths are missing in the 60-69 age group, and at least 500-1000 deaths are missing in the 70-79 and the 80+ age groups.

The logical conclusion of this exercise would be to estimate Delhi’s age based IFRs and compare these with the other countries and regions in this analysis. This is easily done using the estimated death distribution by age groups and the estimated population distributions (both 2011 and 2020). Figure 6 shows that the estimated age based IFRs for Delhi have the same trend as other regions. A rough thumb rule is that IFR increases by a factor of 3 every 10 years as is obvious from the log IFR plot. It is likely that there might be under-reporting of deaths across all age groups in Delhi, and this would show up in the log IFR plot as an upward shift of Delhi’s IFR curve. But, Delhi IFR curve diverges sharply from the trend of all regions above the age of 60 and this can only be explained by a higher share of missing deaths in the records.

**Figure 6:**
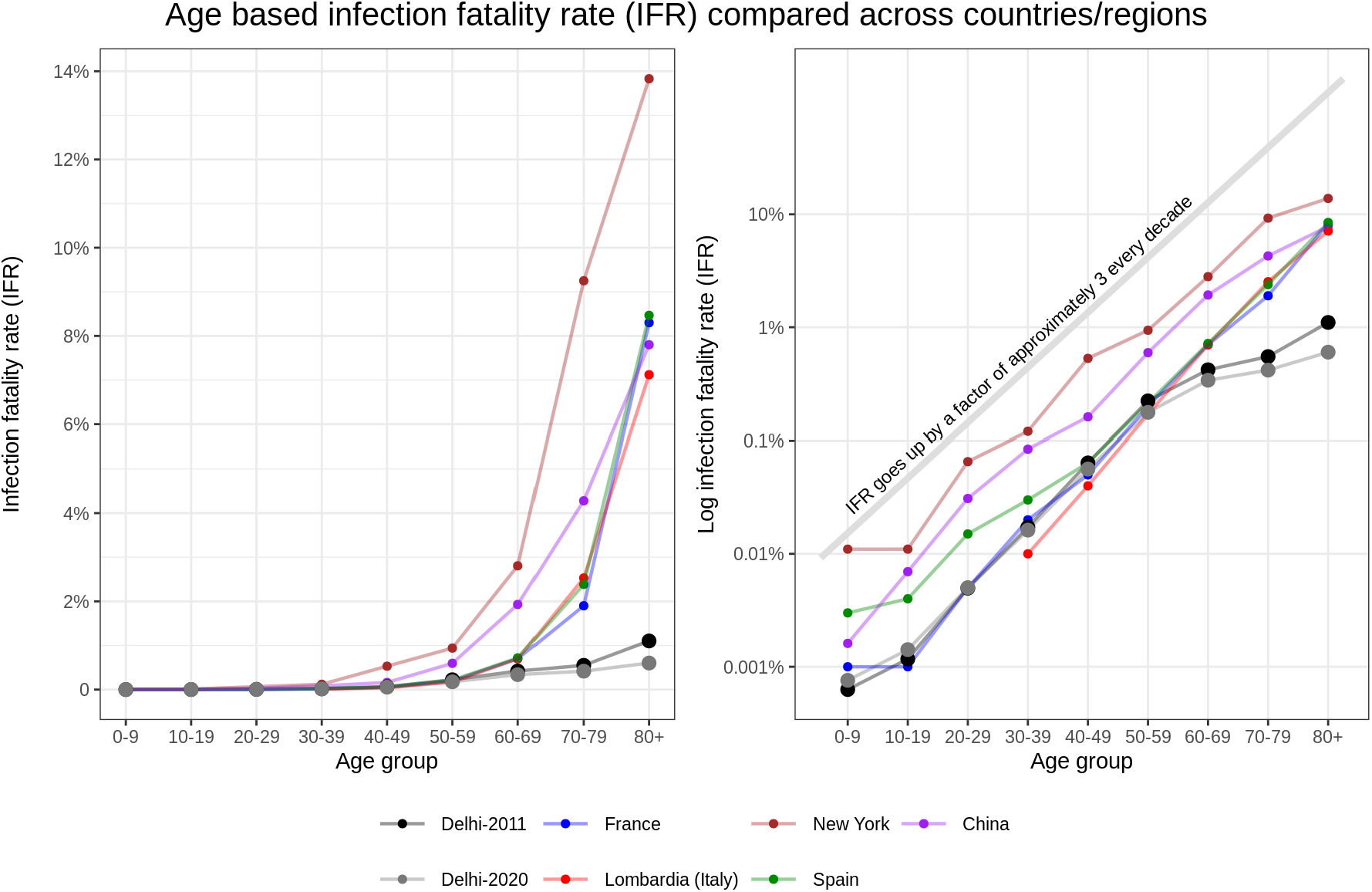
Delhi’s age based infection fatality rate for both 2011 and 2020 population distributions are similar. The IFRs has a same trend when compared to other countries/regions for age groups less than 60. This is easily observed in the log IFR plot, almost all regions fit the thumb rule that the IFR goes up approximately by a factor of 3 per decade. Delhi’s IFRs are significantly lower for 60+ age groups, very likely a result of missing deaths in the records.

## Conclusion

This analysis throws up many puzzling questions regarding the Delhi sero-positivity survey and India’s COVID-19 death records. Before I proceed to list these puzzles and possible resolutions, I would like to reiterate that the assumptions of a) equal infection rate among all age groups, and b) similar death distribution in large urban areas were key to this analysis.

### Is the sero-prevalence an over-estimate?

There is no information in the public domain regarding the survey designs of any of the sero-surveys conducted in the country making it impossible to analyze this issue properly. Given the high prevalence detected in Delhi, it is unlikely that test sensitivity (Pulla 2020a) should be an issue unless the test specificity turns out to be significantly lower than the initial figure of 97.9%. A recent study suggests that Dengue antibodies can give false positives (Nath et al. 2020) in some COVID-19 test kits though it has been suggested that ELISA test kits minimize this possibility (Spinnicci et al. 2020). These concerns should be promptly addressed for future sero-prevalence surveys. One possible resolution of the lower IFR in higher age groups would be that the infection rate and sero-prevalence in these age groups is lower than average. It is important that sero-prevalence surveys are designed to provide estimates of infection rates in different age groups as well.

### Are there any missing or suppressed COVID-19 deaths?

This analysis makes a case for missing deaths in COVID-19 records in India, especially in the 60+ age groups. While this analysis was for Delhi, the death distribution was estimated using Mumbai’s death distribution data (BMC 2020) which was shown to be similar to that of many large urban areas and the national death distribution reported by the Integrated Disease Surveillance Programme (Ghosh 2020). This suggests that missing deaths in the 60+ age group is a nation wide problem. While Mumbai reported lower death registration in March and April (Debroy 2020) in 2020 (compared to 2019), the situation reversed in May 2020 (Slater and Masih 2020), and Mumbai currently reports an excess of about 3000 deaths in January-May 2020 (compared to 2019). The number of COVID-19 deaths till May 2020 was 1279 only, it is likely that some missing COVID-19 deaths form part of the excess deaths reported in May.

A large number of COVID-19 deaths are missed as these deaths might have happened in homes, and it likely that household members were unaware of possibility that these could be COVID-19 related. In some cases, there might have been some symptoms but given the stigma associated with COVID-19, the dead were not tested for SARS-CoV-2 (Rukmini 2020). Delhi and many other parts of the country are also not counting suspected COVID-19 deaths even if funerals were conducted under recommended COVID-19 protocols. Suspected COVID-19 deaths are often not counted as state and city administrations prefer to reduce growth of deaths to show the success of their COVID-19 containment strategies. In some cases, even COVID-19 positive deaths have been classified as deaths under the associated comorbidities (Pulla 2020b). Most large urban areas and a number of states has complete death registration even if they do not have medical certificates on cause of death. These missing COVID-19 deaths are likely to lead to excess deaths that will be reported at a later date.

Timely and accurate data, transparency, and clearly laid out data sharing protocols are an essential public good during an epidemic. It is needless to add that these can save lives, and inform future COVID-19 containment strategies.

## Data Availability

Data available on request

## Notes

### Competing Interest Statement

The authors have declared no competing interest.

### Funding Statement

No external funding was received.

### Author Declarations

Only publicly available data was used so no approval is needed.

### Summary of Updates

Added a disclaimer.

